# Dengue-2 Cosmopolitan genotype detection and emergence in South America

**DOI:** 10.1101/2022.03.29.22273028

**Authors:** Marta Giovanetti, Luiz Augusto Pereira, Gilberto A. Santiago, Vagner Fonseca, Maria Paquita García Mendoza, Carla de Oliveira, Laise de Moraes, Joilson Xavier, Stephane Tosta, Hegger Fristch, Emerson de Castro Barbosa, Evandra Strazza Rodrigues, Dana Figueroa-Romero, Carlos Padilla-Rojas, Omar Cáceres-Rey, Ana Flávia Mendonça, Fernanda de Bruycker Nogueira, Rivaldo Venancio da Cunha, Ana Maria Bispo de Filippis, Carla Freitas, Cassio Roberto Leonel Peterka, Carlos Frederico Campelo de Albuquerque, Leticia Franco, Jairo Andrés Méndez Rico, Jorge L. Muñoz-Jordán, Vinícius Lemes da Silva, Luiz Carlos Junior Alcantara

**Author notes:** Denotes equal contribution (Jorge L. Muñoz-Jordán, Vinícius Lemes da Silva, Luiz Carlos Junior Alcantara). **Correspondence to:** Luiz Carlos Júnior Alcantara.

## Abstract

We report the first confirmed case of DENV-2 Cosmopolitan genotype isolated from a male patient from Aparecida de Goiania, Goías state, Midwest Brazil. Using nanopore sequencing, and phylogenetic analyses our findings provide the first preliminary insight regarding the introduction of this emerging genotype in Brazil and South America.

**Article Summary Line:** Molecular detection of Dengue virus type 2 Cosmopolitan genotype in South America

## Text

Dengue virus (DENV), a single-stranded positive-sense RNA virus with a genome of ∼11,000 kb, is a flavivirus belonging to the *Flavivivridae* family (genus *Flavivirus*) that is transmitted by *Aedes aegypti* and *Ae. albopictus* mosquitos (1). It has caused significant burden and numerous epidemics of mild and severe dengue in the Americas, particularly during recent decades (1). Its diversity can be divided into four antigenically distinct DENV serotypes (DENV-1-4), presenting an inter-serotype nucleotide variability of approximately 30% (2). Each serotype is further subdivided into several phylogenetically distinct genotypes often named according to their geographic origin or other nomenclature, even though some of them have spread to other regions in the past few decades (2). During the past epidemics, DENV outbreaks in Brazil and South America were mainly driven by the circulation of DENV-1-2 serotypes (3,4). Among them, DENV-2 contributes substantially to the dengue burden and dengue-related mortality in the region. DENV-2 includes five distinct non-sylvatic genotypes. Circulation of the Asian I and Asian II genotypes (also labelled as DENV-2 genotype IV) is mostly circumscribed to the Asian continent. The Asian/American genotype, also known as Southeast Asian-American or genotype III, replaced the American genotype (also known as DENV-2 genotype I) in the 80’s (5). The Cosmopolitan genotype (also known as DENV-2 genotype II), is the most wide-spread and genetically heterogeneous genotype, currently circulating in Asia, Middle East, Pacific, and Africa contributing substantially to the dengue burden globally (6). The global dispersal of this genotype may have driven the extensive intra-genotypic diversity, potentially favoring the capacity for wide-spread expansion (6). Cosmopolitan lineages continue to expand geographically, with recent new introductions reported globally, such as in Asia and Africa (7,8). In the Americas, Cosmopolitan genotype reached Peru at the beginning of 2019, spreading mainly along the year in Madre de Dios province, were 4,893 total dengue cases were reported in 2019 (9). However, after its first detection, much is still unknown about its genomic diversity, evolution and transmission dynamics in the region. Since each genotype might be able to lead to a differential clinical outcome and an enhanced virus dispersal, surveillance of circulating strains is pivotal to public health preparedness (5).

Here we report the first confirmed case of DENV-2 Cosmopolitan genotype in Goías state, a well-connected city located in the Midwestern Brazilian region. We further combined mobile genomic sequencing and phylogenetics to provide the first preliminary insights regarding the transmission dynamics of this novel genotype to the Americas. Considering the potential of this genotype to spread within the region, we advocate for a shift to active surveillance to ensure adequate control of any potential outbreak of this genotype across the Americas.

### The Study

A male patient, from the Municipality of Aparecida de Goiania, located in the state of Goiás, Midwest Brazil, presented symptoms (fever, myalgia, nausea retroorbital pain, back pain headache) compatible with arbovirus infections in late November, 2021. A serum sample was collected for molecular diagnosis and sent to the respective Central Public Health Laboratory of Goiás (GO), for molecular screening. Viral RNA was extracted using the QIAmp Viral RNA Mini Kit (Qiagen) and tested by RT-qPCR for arboviral detection (including ZIKV, CHIKV, DENV1-4 and YFV). Molecular testing confirmed DENV-2 infection. To quickly identify the DENV genotype, as part of an active arboviral real-time monitoring effort in collaboration with the Brazilian public health laboratories, genome sequencing was conducted using the Nanopore technology. Briefly, viral RNA was submitted to a cDNA synthesis protocol using ProtoScript II First Strand cDNA Synthesis Kit (10). Then, a multiplex tiling PCR was conducted using Q5 High Fidelity Hot-Start DNA Polymerase (New England Biolabs) and a DENV-2 sequencing primers scheme (3). Amplicons were purified using 1x AMPure XP Beads and cleaned-up PCR product concentrations were measured using Qubit dsDNA HS Assay Kit on a Qubit 3.0 fluorimeter (ThermoFisher). DNA library preparation was carried out using the Ligation Sequencing Kit and the Native Barcoding Kit (NBD104, Oxford Nanopore Technologies, Oxford, UK) (10). The purified PCR products were quantified and DNA concentrations were normalized before barcoding reactions. Sequencing library was loaded onto a R9.4 flow cell, and data were collected for up to 6 hours. Raw files were basecalled using Guppy and barcode demultiplexing was performed using qcat software. Consensus sequences were generated by *de novo* assembly using Genome Detective (https://www.genomedetective.com/) (11). A total of 32,315 mapped reads were obtained, resulting in a sequencing mean depth >1,000X and a coverage of >86%. The DENV-2 Cosmopolitan genotype was identified. The newly sequence data obtained in this study was deposited in GenBank, accession number OM744143.

To put the newly DENV-2 Cosmopolitan genotype sequence in a global context, we constructed phylogenetic trees to explore the relationship of the sequenced genome to those of other isolates of this genotype. We retrieved 1,087 DENV-2 genome sequences from this genotype with associated lineage date and country of collection from GenBank, including the first 2 South American strains isolated in Peru in 2019 (9). Sequences were aligned using MAFFT (12) and edited using AliView (13). Those datasets were assessed for the presence of phylogenetic signal by applying the likelihood mapping analysis implemented in the IQ-TREE 2 software (14). A maximum likelihood phylogeny was reconstructed using IQ-TREE 2 software under the HKY+G4 substitution model (14). We inferred time-scaled trees by using TreeTime (15).

Our phylogenetic analysis estimated by the DENV typing tool (available at http://genomedetective.com/app/typingtool/dengue), consistently place the Brazilian strain in a clade within the Cosmopolitan lineage with maximum statistical support (bootstrap = 100%) (**Figure S1**). Time-resolved maximum likelihood trees showed that the new isolate obtained in this study clusters with two recently described DENV-2 strains isolated in Peru in 2019 (strong bootstrap support of 96%) (**Figure 1 A-C**), suggesting a possible cross-border transmission. This cluster containing the South American isolates diverges from strains from Bangladesh collected between 2017-2019 (bootstrap support 1.0), suggesting a complex transmission scenario of introduction events likely mediated by trans-continental travel (**Figure 1 A-C**). Additional sampling of the region is needed to accurately reconstruct the routes of importation and transmission dynamics into Brazil and South America and the epidemiological impact upon the spread of the Cosmopolitan genotype.

**Figure 1.**
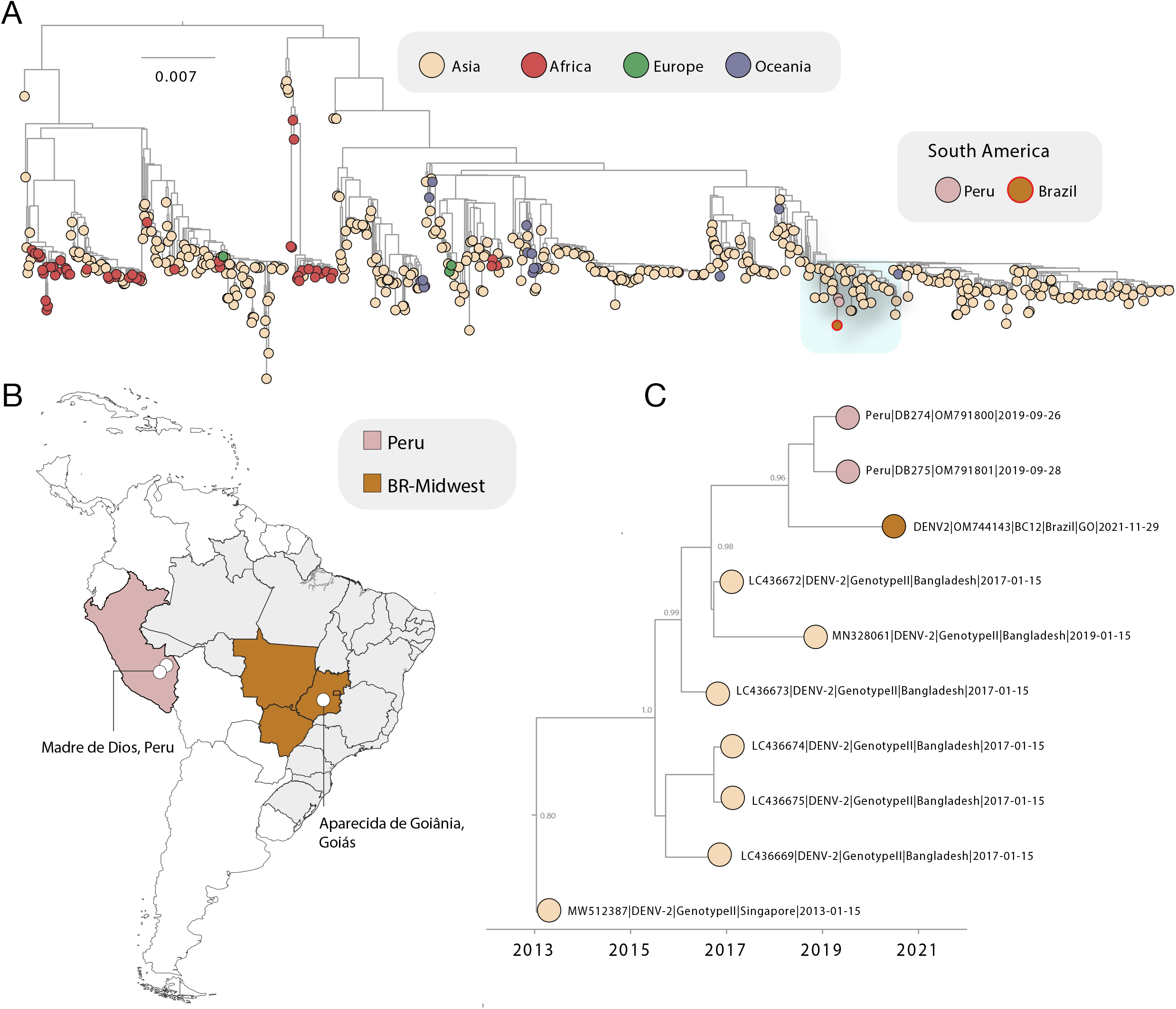
Investigation of DENV-2 Cosmopolitan genotype in Brazil. A) Maximum likelihood (ML) phylogenetic analysis including the newly complete genome sequence from DENV-2 Cosmopolitan genotype generated in this study plus 1,089 publicly available sequences from GenBank. The scale bar is in units of nucleotide substitutions per site (s/s) and the tree is mid-pointed rooted. Colors represent different sampling locations. B) Map of Brazil and Americas showing sampling location of the first DENV-2 Cosmopolitan genotype genome reported in Brazil and characterized in this study (white circles); C) Time-resolved maximum likelihood tree showing the DENV-2 Cosmopolitan cluster containing the newly genome generated in this study. Colors indicate geographic location of sampling. Support for branching structure is shown by bootstrap values at key nodes.

## Conclusions

The emergence of the DENV-2 Cosmopolitan genotype in the Americas merits active outbreak risk assessment and surveillance across the region to prevent further spread and reduce epidemic potential.

## Data Availability

The newly sequence data obtained in this study was deposited in GenBank, accession number OM744143.

## Ethics statement

This project was reviewed and approved by the Pan American Health Organization Ethics Review Committee (PAHOERC) (Ref. No. PAHO-2016-08-0029) and by the Federal University of Minas Gerais (CEP/CAAE: 32912820.6.1001.5149). The availability of these samples for research purposes during outbreaks of national concern is allowed to the terms of the 510/2016 Resolution of the National Ethical Committee for Research – Brazilian Ministry of Health (CONEP - Comissão Nacional de Ética em Pesquisa, Ministério da Saúde), that authorize, without the necessity of an informed consent, the use of clinical samples collected in the Brazilian Central Public Health Laboratories to accelerate knowledge building and contribute to surveillance and outbreak response. The sample processed in this study was obtained anonymously from material exceeding the routine diagnosis of arboviruses in Brazilian public health laboratories that belong to the public network within BrMoH.

## Biographical sketch

Dr. Giovanetti is a Visiting Researcher at the Reference laboratory of Flavivirus, Fiocruz Rio de Janeiro, Southeast Brazil. Her research focuses on investigating the patterns of gene flow in pathogen populations, focusing in phylogenetics and phylogeography as tools to recreate and understand the determinants of viral outbreaks and how this information can be translated into public policy recommendations.

## Acknowledgement

This work was support in part through National Institutes of Health USA grant U01 AI151698 for the United World Antiviral Research Network (UWARN), the National Council for Scientific Development- CNPq (421598/2018-2) and by the Brazilian Ministry of Health (SCON2021-00180). MG and LCJA are supported by Fundação de Amparo a Pesquisa do Estado do Rio de Janeiro (FAPERJ). Peru DENV 2 sequences were obtained in the frame of the PAHO VIGENDA project (<u>VI</u>gilancia <u>GE</u>nomica de <u>D</u>engue en las <u>A</u>mericas as per is acronym in Spanish).

## Author contributions

Conception and design: MG; and LCJA; Investigations: MG; LAP; GAS; VF; MPGM; CdO; LdM; JX; ST; HF; EdCB; ESR; DFR; CPR; OCR; ANM; FdBN; Data Analysis: MG; VF; LdM; and GB;

Writing – Original: MG and LCJA; Draft Preparation: MG; GAS; MPGM; LF; JAMMR; JLMJ; and LCJA; Revision: MG; LAP; GAS; VF; MPGM; CdO; LdM; JX; ST; HF; EdCB; ESR; DFR; CPR; OCR; AFM; FdBN; RVdC; AMBdF; CF; CRLP; CFCdA; LF; JAMR; JLMJ; VLdS; and LCJA; Resources: LAP; AFM; MPGM; AMBdF; RVdC; VLdS, CFCdA; CRLP; CF; JAMMR; JLMJ; and LCJA.

## Supporting information

**Supplementary Figure 1.**
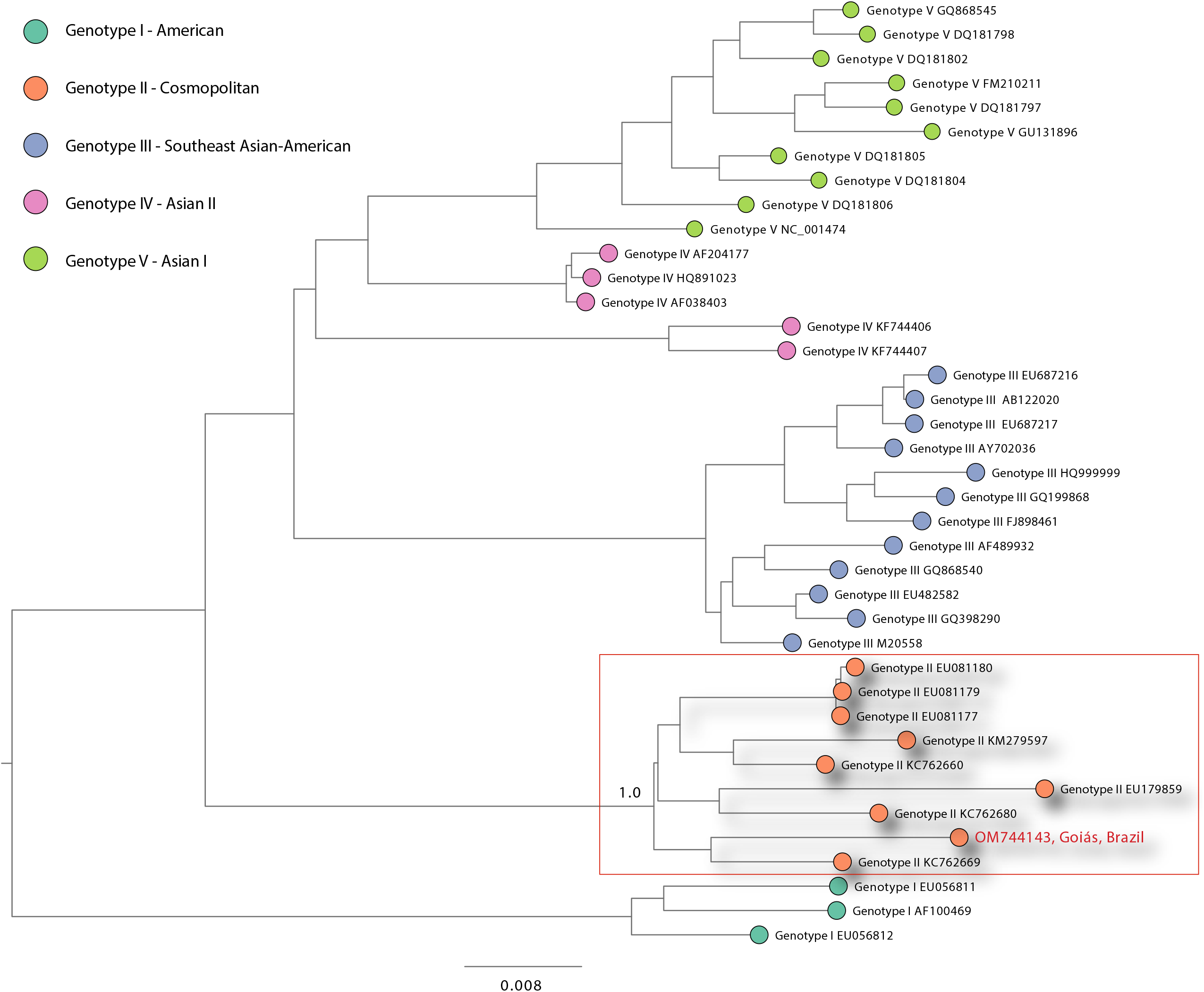
DENV-2 genotyping assessment. Maximum Likelihood phylogeny comprising the newly genome sequence (highlighted in red) generated in this study plus 37 sequences retrieved from NCBI belonging to all DENV-2 non-sylvatic genotypes (DENV-2 Genotype I – American; DENV-2 Genotype II – Cosmopolitan; DENV-2 Genotype III - Southern Asian-American; DENV-2 Genotype IV - Asian II; DENV-2 Genotype V - Asian I). Dengue 2 virus genotypes are coloured according to the legend.

